# EWA-DB, Slovak Database of Speech Affected by Neurodegenerative Diseases

**DOI:** 10.1101/2023.10.13.23296810

**Authors:** Milan Rusko, Róbert Sabo, Marián Trnka, Alfréd Zimmermann, Richard Malaschitz, Eugen Ružický, Petra Brandoburová, Viktória Kevická, Matej Škorvánek

## Abstract

A new Slovak speech database EWA-DB was created for research purposes aimed at early detection of neurodegenerative diseases from speech. It contains 1649 speakers performing various speech and language tasks, such as sustained vowel phonation, diadochokinesis, naming and picture description. The sample of speakers consists of individuals with Alzheimer’s disease, mild cognitive impairment, Parkinson’s disease, and healthy controls. In this article we describe the EWA-DB development process, the language and speech task selection, patient and healthy control recruitment, as well as the testing and recording protocol. The structure and content of the database and file formats are described in detail. We assume that the presented database could be suitable for the development of automatic systems predicting the diagnoses of Alzheimer’s disease, mild cognitive impairment, and Parkinson’s disease from language and speech features.

## Background & Summary

Alzheimer’s disease (AD), and Parkinson’s disease (PD), are the two most common neurodegenerative diseases, which have been historically diagnosed based on their clinical symptoms, however, additional diagnostic tools such as imaging or biological markers are being increasingly implemented in their clinical diagnostic criteria^1, 2^. While these diagnostic criteria require specific clinical expertise or availability of more sophisticated investigation methods, the need for early (even prodromal) diagnosis and screening of subjects at risk is becoming increasingly important. In this regard, mild cognitive impairment seems to be the most relevant risk factor for development of fully manifested Alzheimer’s disease, and updated research criteria for prodromal Parkinson’s disease have been also published recently^3, 4^.

According to the World Health Organization more than 55 million people worldwide suffer from AD or other forms of dementia and there are nearly 10 million new cases every year^5^. The patient’s speech and language are commonly affected by these disorders and specific changes can be observed even in their prodromal phases^6^, which can be utilized in disease diagnostics and further monitoring.

A review of works on speech analysis of people with neurodegenerative diseases up to 2017 can be found in the work of Boschi et al.^7^.

As summed up by Chen^8^ assessment of cognitive function is typically carried out by a trained psychometrician or neuropsychologist using a battery of cognitive tests that examine various aspects of cognitive abilities, including language skills^9, 10^. Neuropsychological testing is a time-consuming process that can take up to several hours. Also, in many cases repeated assessments are necessary to monitor the progression of cognitive decline. There is currently a growing interest in creating automated assessment methods that will speed up the process of early detection of people with neurodegenerative diseases based on their cognitive and especially language difficulties^8^.

As for speech and language, Alzheimer’s disease is most pronounced at the lexical-semantic, discourse-pragmatic, syntactic and phonetic levels^7^. Studies using machine learning techniques for speech analysis for MCI and AD patients show that different parameters have different weights for the diagnosis of AD and MCI and that the combination of several parameters improves the accuracy of neurodegenerative disease prediction^11^. In Parkinson’s disease, motor speech problems in the sense of hypokinetic dysarthria prevail, which affect several subsystems of speech - phonation, resonance, articulation and prosody^12^ and are captured by parameters such as pause duration^13^ and prosody change^14^. Symptoms of PD are present not only in speech but also in language, mainly in morpho-syntactic processing, as in verb processing^15^.

The existence and availability of a speech database that represents the investigated phenomena on a sufficient sample of patients with neurodegenerative diseases and healthy people is a necessary condition for research into the possibilities of using automatic speech processing and machine learning methods to predict neurodegenerative diseases from speech.

For the two most widespread neurodegenerative diseases, PD and AD, there are several types of speech databases in the world. Some of them are published and accessible in the form of corpus description, fewer of them are published in the form of recordings. Some of them are accessible on request and the smallest group are freely accessible PD^16^ and AD^17^ databases that can be directly used for machine learning.

Furthermore, we can divide speech databases into groups according to the type of tasks that are recorded during its creation. The most widespread types of tasks that the databases contain are for PD^18, 19^: phonation of sustained vowels, reading and repetition of sentences, description of pictures or diadochokinesis tasks (e.g., rapid repetition of syllable sequence /pataka/); and in patients with AD^20, 21^: picture description, fluent questions answering, object naming or narratives on the topic of daily life.

The most frequent problems associated with this type of databases are inconsistency across datasets, extremely specific purpose of use, poor quality of recordings or dataset imbalance^22,23^. Another problem is the small number of recordings in the dataset. In this case, there is not enough data for training and the machine learning techniques cannot show their full potential. Databases are also language specific, and it is problematic to find different datasets which can be combined.

Language specificity creates perhaps the biggest challenge. To name an example, phonemic diversity in different languages represents a significant challenge for the evaluation of speech/language and their automatic processing through machine learning and ultimately for the final prediction of neurodegenerative diseases^24^. For instance, Anglo-Germanic languages are characterized by frequent clusters of consonants, while in Roman languages the consonant-vowel structure predominates^25^. Due to language differences and the challenges, they can generate, we find the creation of new language specific databases necessary.

This manuscript presents a new Slovak speech database created for AD, MCI, and PD research and for development of automatic systems predicting these diagnoses from language and speech features.

## Methods

As manifested in the previous chapter, creation of language specific databases is needed. When creating the Slovak database, we followed the current International Parkinson and Movement Disorder Society Guidelines for Speech Recording and Acoustic Analyses in Dysarthrias of Movement Disorders^24^. We also considered the language and speech testing tasks, which are used in common clinical practice and at the same time are shown by research to be suitable materials for obtaining a speech sample for its automatic analysis. As for the participants, we considered it important to represent the most common neurodegenerative diseases, which are PD and AD/MCI, as well as the healthy population for the purposes of creating the possibility of comparison with normative performances.

### Selection and compilation of speech and language tasks

Speech and language tasks that are known to be sensitive to capture early changes in the speech and language production of PD, AD and MCI patients were selected.

### Sustained vowel phonation and diadochokinesis

Since a high percentage of PD patients have voice difficulties related to dysarthria, it is crucial to include tests that evaluate these symptoms. Sustained vowel phonation and sequential and alternating movement tasks, such as diadochokinesis, are among the basic voice tasks included in the voice diagnosis of patients with PD.

For the purposes of this project, following the guidelines by Rusz, participants were first instructed to take a deep breath and perform a sustained phonation of vowel /a/ as long and steadily until they run out of air or until the end of recording, which was set at 15 seconds. The ideal minimum duration of phonation recommended for hypokinetic dysarthria patients, including PD, is 6 seconds^24^.

Diadochokinesis is a dysarthria evaluation method commonly applied in clinical practice, which tells about the ability of the maximum speed of syllable repetition using alternating or sequential movement tasks. The alternating task measures the rapid repetition of a single syllable, while the sequential task measures the rapid repetition of syllable sequences^26^.

In our project, the sequential motion rate was included. Participants were instructed to take a deep breath and repeat syllable sequence /pataka/ as quickly and accurately as possible until told to stop. The instruction was to pronounce continuously, intelligibly and to speak as quickly as they can without being imprecise^27^. The duration of this test recording was set at 8 seconds to allow for at least 12 sequence repetitions performed with one breath as suggested by the above-mentioned guidelines^24^.

### Object and action naming

Confrontational naming relies on specific brain networks, and involves various cognitive processes such as visual recognition, semantic activation, lexical retrieval and articulation^28, 29^. In the task of confrontational naming, perceptual difficulties, visual perception errors, impaired phonological and semantic access is documented in patients with AD^30^. Difficulties in picture naming are also documented in PD^31^.

There are several visual confrontation naming measures available. The most often used confrontational naming test is the Boston Naming Test (BNT). It is commonly used by neuropsychologists and speech therapists to assess lexical retrieval. The original version includes 85 items as simple line drawn pictures^32^ and it was later shortened to 60 items^33^. The Slovak version of the test is not available so far. An original Slovak picture naming test is available^34^, containing black-and-white drawings of 30 objects and 30 actions. However, this test appears to be insufficiently sensitive to discrete language disruptions as those manifested in early PD^35^.

In the design of the visual confrontation naming task for EWA-DB our own set of 30 object and 30 action photographs was created. Choosing the format for full colour was based on the work of Li et al.^29^ which describes several reasons for coloured image preferences, such as criticism of the black-and-white line drawings in the BNT^36^, possible cohort effects and cultural bias that make these drawings liable to be misperceived in non-English speaking population^37^, or a questionable diagnostic validity^38^. In comparison, a meta-analysis of studies aimed at picture naming showed improvement in naming accuracy and response times when using coloured images^39^. In the study of Li et al.^29^ the original version of the BNT and a colour version of BNT were compared. The study documented higher scores in naming accuracy and an overall better diagnostic accuracy for MCI and AD detection when the coloured version of the test was used^29^.

In addition to colour, another factor affecting picture naming ability is the nature of the objects in the sense of biological and artifact items. When the ability to name biological and artifact items was observed, individuals with MCI showed lower performance on biological items compared to cognitively healthy individuals, suggesting a category-specific impairment of biological items in MCI^40, 41, 42, 43^.

Not only objects but also actions are part of the naming test created for EWA-DB. The reason is that naming processes for verbs and nouns are different. As explained by Hwang et al.^44^, verbs and nouns belong to separate grammatical classes and the difference is also in terms of their semantic representation as nouns refer to objects, while verbs refer to actions. Nouns and verbs engage different brain areas. Lexical retrieval of nouns is engaging left temporal areas and verb retrieval is engaging left frontal areas^44, 45^. This could be one of possible explanations for difficulties of verb production in PD. Several studies of PD document action naming deficits, thought to reflect the presence of frontal and prefrontal dysfunction^46^, namely in the areas of pre- and post-central gyri bilaterally, left frontal operculum, left supplementary motor area^47^.

In the final picture set for EWA-DB 30 nouns and 30 verbs were selected. Their selection was based on the criteria of age of acquisition, frequency, and word length. Some additional criteria for the visual form of stimuli were added:

- The number of high-frequency words is ⅓ compared to low-frequency words. The frequency is determined based on information from the Slovak National Corpus. For a word to be considered high frequency, it’s occurrence in Slovak language had to be greater than 1000.
- The pictures had to be as simple as possible so that there is no doubt about what the main object or main activity of the image is. Images without a background were chosen to focus attention on the object itself.
- The pictures could not be too detailed as the pictures were planned to be presented by smartphone screens and given their age, we assumed that some users may have vision problems.
- The pictures had to be square or vertically oriented rectangle, so that their details can be recognized well on the smartphone screen in a vertical position.
- The pictures are intended for Slovak speakers, mainly in the age of 50-80. Objects and activities were chosen that were well known and frequently encountered by these persons during their youth and active adulthood. For example, a soccer ball would be selected for a ball image, rather than an American football ball.

The pictures are included in the database in a readme.txt file.

### Picture description

The analysis of spontaneous speech has been receiving more and more attention. In AD, changes in lexis, grammar, informativeness^48^, cohesion and coherence are documented, while the most evident are the changes in fluency and semantics^49^. Since already in MCI and early AD disruptions of the temporal parameters of speech can be found (speech tempo, number of pauses and their length) the computerized analysis of spontaneous speech may be a promising tool for early AD detection^50^. Disruptions of spontaneous speech are also captured in PD. The spontaneous speech of patients with PD is less fluent, monotonous, and syntactically simpler with a disturbance in the informativeness present^51^. In recent years, attention has also been paid to cohesion and coherence in early PD. A higher number of incorrect connective ties and thus a lower degree of cohesion is documented^52^. Difficulties in this area are present not only in cohesion, but also in coherence, namely global coherence^53^.

There are several ways to obtain a sample of spontaneous speech, such as interviewing, storytelling or picture description. Spontaneous language analysis with a picture description task is useful to detect subtle language impairments even in early stages of AD^54^. Also in PD, spontaneous speech assessment is considered to have the best ability to differentiate PD patients from healthy adults^55^.

The most widely used task in neurodegenerative diseases is the Cookie Theft from the Boston Aphasia Diagnostic Examination^56^. This task has proven to be an effective tool in several neurogenic disorders, including AD and MCI^57^. However, for the purposes of our project, we encountered several difficulties in implementing this image, such as its landscape orientation not being suitable for mobile phone displays or the inscription being in English. We also wanted to remain consistent when creating images for the database, so we preferred colour images over black and white. We expect the advantages of colour images to be the same as we describe in the section on creating images for the naming task. We therefore created new images for this project, while we tried to incorporate those features of the Cookie Theft picture, which make it such an effective tool, as described by Cummings^58^. The pictures we created contain people, objects, activities and situations, the description of which can best capture the differences between healthy expression and the expression of a person with beginning AD or PD. According to Cummings^58^, there are 7 areas assessed in picture description. Namely points 2 to 5 had to be considered in the design of our images by incorporating people, objects and activities containing areas and categories that are important for the assessment of our patient’s speech. The areas of interest are as follows:

- **Significant features, importance of information.** The patient should present a clear distinction between essential and minor matters; the chronology of his statement should be correct in the sense of starting with the essentials.
- **Semantic categories.** AD patients show a tendency to use more general, less specific terms e.g., *woman* is more general than *mother*. The picture also must depict actions e.g., *the boy falls*.
- **Referential cohesion.** The use of deixis (*he*, *she*) in a way that it is clear which person or object the deixis represents, which is a problem for AD and PD patients.
- **Causal and temporal relations.** The patient should describe causalities - that the water flows out because the woman left the tap open; the children steal biscuits because they saw that the mother was not looking etc. In AD the context is often incomprehensible.
- **The language of mental state.** Cognitive mental state includes knowledge, beliefs, and assumptions. Affective mental state is represented by emotions such as happiness or anger - the events and actions in the picture can only be explained by the mental state of the persons. Typical descriptive words of the language of the mental state are for instance *wants*, *dreamy*, *careful*, *forgot about* etc. In AD they are often missing.
- **Language structure and speech.** Assessment of language and motor speech structure skills, such as word search, pauses, auxiliary sounds, replacement of words with a more general form (e.g., *that*, *stuff*, *someone*, *person*), neologisms and other phonological errors.
- **General knowledge and perception.** Some defects in general knowledge and perception, such as forgetting that one has already described something and describes one scene several times or giving a description of only one of the sides of the image (right or left). However, this occurs rather in patients with severe dementia^58^.

Based on the described criteria, five pictures were created for the purposes of the EWA-DB – two simple black-and-white pictures and three complex coloured pictures. An example of these pictures is shown in Fig. 1.

**Fig. 1.**
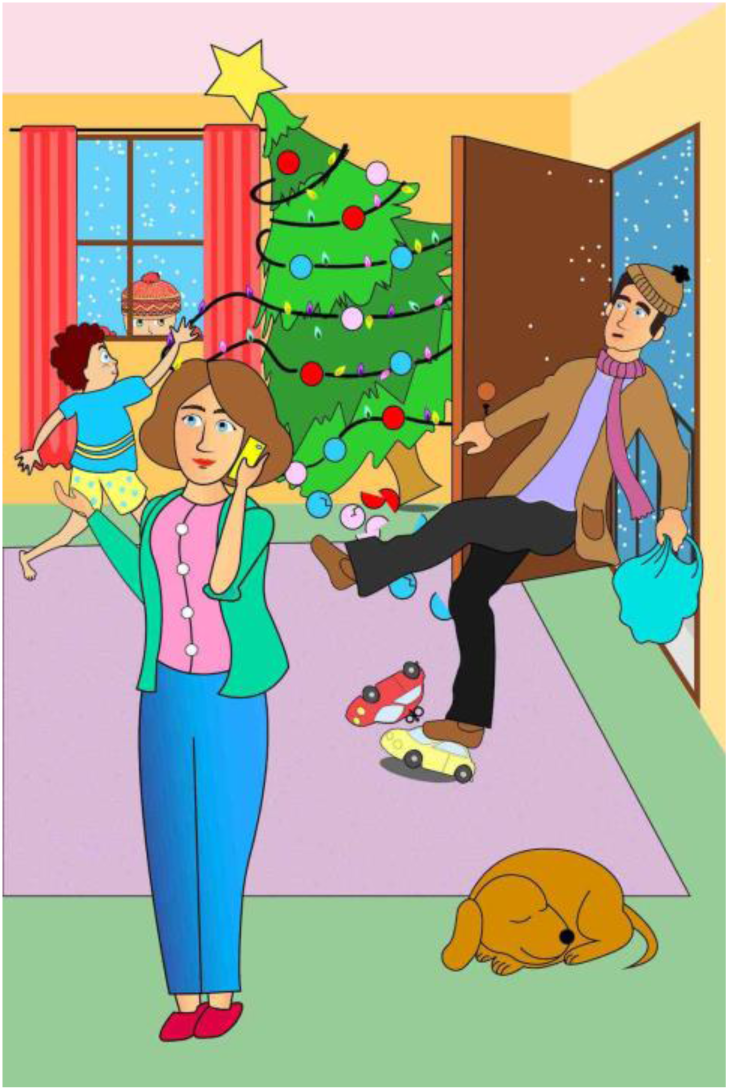
Example of a picture used in the picture description task.

### Recruitment of participants

Since the goal of this project was a new Slovak speech database created for AD, MCI and PD research, the recruitment of such patients along with healthy controls took place.

To recruit a large number of participants from all regions of Slovakia, we reported on the ongoing project in newspapers, on the radio, and on television. We distributed leaflets mainly to retirement homes and general practitioners’ clinics. An association of retirement companies provided a database of 140 facilities that were approached with a request for cooperation. From the approached organizations and contacts, the interested parties announced themselves via e-mail or telephone. Subsequently, the interested party was sent an informative article about the details of the research process and a personal meeting was arranged. MCI and AD participants were mainly recruited from the MEMORY Centre - a specialized centre for diagnosis, treatment, and education in the field of memory disorders and dementia. PD participants were mainly recruited from the Department of Neurology and Centre for Rare Movement Disorders of the L. Pasteur University Hospital. So, the group of positively diagnosed patients consists of people who have already been diagnosed with MCI, AD, or PD during a medical examination.

During the recruitment the following methods and examinations were used.

**The anamnestic questionnaire** contains items for obtaining basic descriptive characteristics (gender, age, education, and lifestyle factors). Although the collection of samples of speech recordings is not quota-based, the aim is a heterogeneous proportional representation in relation to the monitored variables.

**The subtest Similarities** from the WAIS-III is considered one of the best instruments for measuring the index of the verbal comprehension factor, in which crystallized intelligence is significantly involved^59, 60, 61^. Premorbid intelligence is a concept through which the impact of neurological damage on cognitive performance is evaluated as an estimate of the basic, “premorbid” global performance before the onset of damage^62^. The total score in the subtest is an indicator of the current level of the examinee’s abstraction ability and overall verbal comprehension. The test is also recommended as a suitable tool for the evaluation of semantics and language from the available tests adapted in our region.

**Montreal Cognitive Assessment** (MoCA)^63^ is a screening tool aimed at assessing the global cognitive status by including the assessment cognitive domains such as executive functions, visuospatial functions, memory, attention, speech, orientation in time and space. The scoring ranges from 0 – 30 points. The test itself states a cut off score of 26 points. However, in older adults, the average performance ranges from 26 ± 3 points and is also dependent on education^64^.

### Inclusion and exclusion criteria

When creating the database and recruitment of participants, we followed these **inclusion and exclusion criteria**:

**MCI.** Classification criteria according to the diagnostic criteria of Albert et al.^65^, age 50-90 years, MoCA score 25-23, preserved activities of daily living measured by Barthel’s Index for Activities of Daily Living, score in the questionnaire GDS ≤ 9 b, GAD-7 ≤ 9 b. Participants had to be diagnosed as patients with MCI by a psychiatrist or a neurologist according to the Albert et al.^65^ criteria for a diagnosis of MCI. Exclusion criteria for the MCI group were as follows: (1) history of or current psychiatric disorder; (2) history or neurological evidence of stroke, head injury, or neurodegenerative disorders that are known to influence cognitive functioning; and (3) on medication for depression and/or Alzheimer’s disease.

**AD.** Diagnosis established according to the criteria of the International Classification of Diseases (ICD-10) or the Diagnostic and Statistical Manual of Mental Disorders (DSM-IV) and in all stages of the disease confirmed by a specialist (psychiatrist or neurologist), age 50-90 years, MoCA score between 22-18, capability to give informed consent.

**PD.** Classification criteria according to MDS criteria^2^ or according to MDS criteria for prodromal Parkinson’s disease^4^, prodromal or early stage of the disease (Hoehn & Yahr stage I or II) and duration of the disease less than 10 years, MoCA score 20 and more, patient examined in the ON state possibly before starting treatment, age over 18.

All participants also met the inclusion criteria: normal or compensated vision and hearing, being a native speaker of Slovak language, no clinical history of head injury or psychosis, no medical record of drugs or alcohol consumption, not being under pharmacological treatment affecting cognitive functions, absence of disorders with expected impact on language and speech.

Exclusion criteria for healthy and clinical groups were the self-reporting of a previous stroke, brain tumours or psychiatric disorders such as bipolar disorder or schizophrenia, current or past alcohol or drug abuse history, or under-corrected auditory or vision difficulties.

The recruited participants were invited to various recording sites throughout Slovakia and underwent the same recording procedure administered by trained staff (administrators). The team of administrators consisted of healthcare workers, such as psychologists, doctors, nurses, and trained staff or students (mainly students of psychology or speech and language pathology).

### Testing and recording procedure

The testing and recording procedure were performed in the following steps. Participants signed a written informed consent prior to participating in the research, including information about personal data processing, providing health status information and sound recordings publishing. The study was undergone in accordance with the Declaration of Helsinki, including the ethics committee approval of the Bratislava self-governing region.

Following that, the anamnestic questionnaire and measures for the inclusion criteria were administered (see above for details). Finally, the sound recording protocol was administered. It consisted of speech and language tasks: sustained vowel phonation, diadochokinesis, object and action naming and picture description, the designs of which were described in detail in the section above.

### Data collection and processing

Commonly available mobile phones were used for data collection. Both Apple and Android devices have been used. At the beginning of the project, older phone models were also tested, and the recording software was modified to use only mono recording on all devices, without the use of compression with 16 bit depth and sampling rate of 16kHz. Audio files were saved in Microsoft waveform format with Pulse Code Modulation.

The recording procedure started with a calibration phase during which the usable screen size was determined, the functionality of the recording and the reliability of automatic speech recognition on the voice of the participant were verified.

A smartphone application was designed that covers the entire process of collecting data from participants:

- Anamnestic questionnaire. Basic information about the participant (age, gender, education, lifestyle factors) is inserted by the administrator. Sensitive data (first name, last name, contact details) are protected by the GDPR law and are not processed electronically. They are collected in writing and archived together with the data processing consent in accordance with the GDPR law.
- Measures for the inclusion criteria (see Inclusion and exclusion criteria).
- Sound recordings of sustained vowel phonation and diadochokinesis.
- Sound recordings of picture naming. The application sequentially displays 60 pictures (30 objects and 30 actions). Participants are instructed to name the object or action using one word. Moving from one picture to another is done by pressing a button. As we discovered at the beginning of the project, people tend to move to the next image too soon, press the button and end the recording before finishing the whole word. Therefore, the application stops recording with a short delay. The standard recording time of one image is 2-3 seconds.
- Sound recordings of picture descriptions. The application presents 2 simple black and white pictures and 3 complex color pictures. Participants were asked to describe these pictures in detail. The recording time was set to 30 seconds for simple pictures and 90 seconds for the complex ones.
- Informed consent for data processing and publication.

A server application was created to collect and process data from mobile phones used in our project. All data from the mobile phones, data supplemented with the date of testing, name of the administrator, and technical information about the version of the application and the mobile phone itself were uploaded to a MySQL database^66^.

A total of 120 answers to questions from the questionnaires used in the inclusion criteria and 65 audio recordings per participant, are stored in a database. It is possible and recommended to record when the mobile is disconnected from the internet and in airplane mode so that the test is not interrupted. The application sends the data to the server when it is connected to the internet. The server uses a MySQL database^66^ to store metadata. The audio data is stored in the Minio^67^ database. Due to the different devices that can be used, recordings are stored in PCM Microsoft wav format with 16 kHz sampling and 16 bit resolution. The server application provides a web interface where all data can be viewed, listened to and audio recordings can be downloaded. The interface also allows to display statistical data, and it is possible to filter the data, e. g. by age, diagnosis, gender, etc. as well as to export some specially selected parameters to an Excel sheet.

To allow further data processing by artificial intelligence, various features were extracted from the audio files:

- Text transcription of speech recordings, with time marks for individual words and long and short pauses.
- Length of the whole recording and duration of the speech itself.
- Number of words, number of syllables.
- Speaking rate in words per minute, syllables per minute.
- Response time.
- Time needed for a correct answer in the naming test.
- Number and types of hesitations in speech.
- Features extracted using Neurospeech toolkit^68^.
- Features extracted using OpenSmile toolkit^69^ with Gemaps settings^70^.
- Trill Embedding^71^. F

### Annotation

To achieve that the recordings can be used for training specific acoustic and language models, collected audio files needed to be transcribed. First, automatic transcriptions were created using a speech recognizer developed at the Institute of Informatics of the Slovak Academy of Sciences. Subsequently, the automatic transcriptions were corrected by trained annotators using the program Transcriber 1.5.1.^72^ and adding labels for different acoustic events.

Hesitations are one of the most frequently observed parameters in spontaneous speech of patients with cognitive decline and/or dementia. However, the term hesitation is not used uniformly throughout various studies focused on spontaneous speech analysis. While in some cases the term hesitation is used in a narrow sense, such as absence of speech during more than 30ms^73^, in other cases it is used as an umbrella term^50^ including various manifestations, as in silent pauses, filled pauses, but also verbal expressions such as false starts or restarts, interjections, or prolonged sounds of words. In our annotation convention, we were primarily guided by whether the hesitation was verbal or non-verbal. Non-verbal, but vocal expressions were tagged as [hez] - typically a schwa sound, but it could also be any other vocal expressions indicating word-finding difficulties. Silent pause tags were not introduced. Non-verbal sounds with a certain meaning, which did not indicate word-finding difficulties, were transcribed verbatim and tagged with a % symbol, e. g. %a: in the meaning of surprise or %hm in the meaning of thinking. Non-verbal sounds without any meaning were marked with a different tag [spk], e. g. in the case of mouth smacking or throat clearing. Of the verbal expressions, we paid attention to false starts and restarts, which were tagged by their joint tag +[ned], that was added to the beginning of the word being falsely started or restarted. Furthermore, we were paying attention to interjections, which were literally transcribed. We also considered sound prolongations to be a form of hesitation, as they can occur as results of word-finding difficulties. Such manifestations were given a separate tag – a colon was written after the sound concerned.

As hypokinetic dysarthria is present in many patients with PD, a reduced degree of speech intelligibility was expected. Words that were less intelligible were transcribed verbatim and placed in two parentheses ((word)). Words that were not intelligible at all were also placed in two parentheses and marked with as many “x” letters as the unintelligible word had syllables, e. g. ((x)) represents a monosyllabic unintelligible word. As changes in respiration are also documented in PD, the [ex] tag was used to indicate prolonged or loud exhalations.

We expected phonetic-phonological difficulties in both patients with PD^74^ and AD^75^. Differences in pronunciation were observed and tagged using the forward slash symbol. First the target pronunciation was written down following the forward slash symbol and the actual pronunciation of the participant concerned. Thus, also slips of the tongue and phonemic or phonetic paraphasia were marked using this tag.

When working with the elderly and especially when collecting data from patients with neurodegenerative diseases, the input of investigators was necessary in some cases, e. g. to refine the instruction. Such entries were manually removed from the transcripts and the segment was marked as [inv].

The last tag included was [ruch]. This marked all background disturbances that could affect the correct capture of analyzed speech.

## Data Records

The EWA-DB speech database is publicly available at ELDA^76^ under the name EWA-DB. The database contains 1649 speakers, of which 87 are AD patients, 175 are PD patients, 2 speakers have a combination of AD and PD, 62 are MCI patients and 1323 are healthy controls (HC). The distribution of the clinical samples and the sample of healthy controls in terms of age, education, and gender (factors that may influence performance on language tests) is shown in Tables 1, 2, and 3.

**Table 1.** Distribution of EWA-DB according to age.

| AGE |  |  |  |  |  |  |
| --- | --- | --- | --- | --- | --- | --- |
|  | AD | AD-PD | HC | MCI | PD | SUM |
| <50 | - | - | 66 | - | 13 | **79** |
| 50-59 | - | - | 341 | - | 22 | **363** |
| 60-69 | 6 | 1 | 459 | 6 | 62 | **534** |
| 70-79 | 35 | 1 | 320 | 32 | 54 | **442** |
| >=80 | 45 | - | 135 | 23 | 24 | **227** |
| missing data | 1 | - | 2 | 1 | - | **4** |
| **SUM** | **87** | **2** | **1323** | **62** | **175** |  |

**Table 2.**
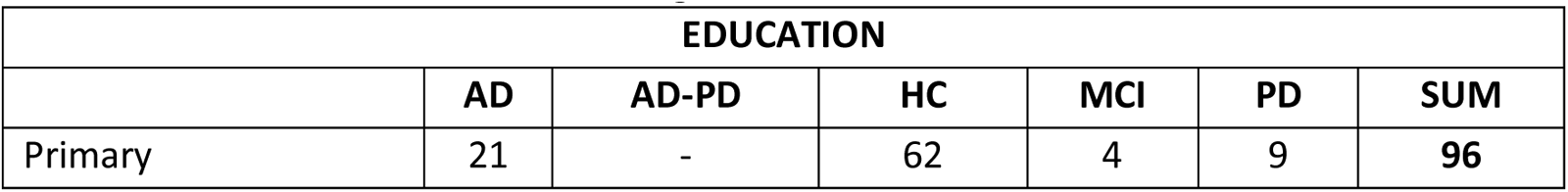
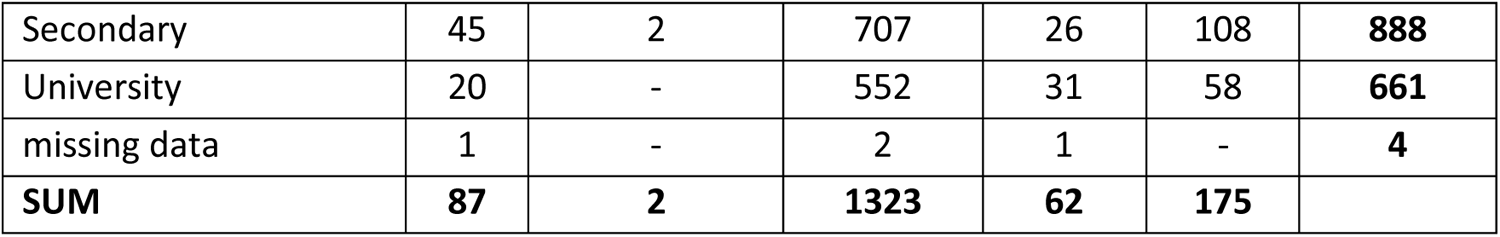
Distribution of EWA-DB according to education.

**Table 3.**
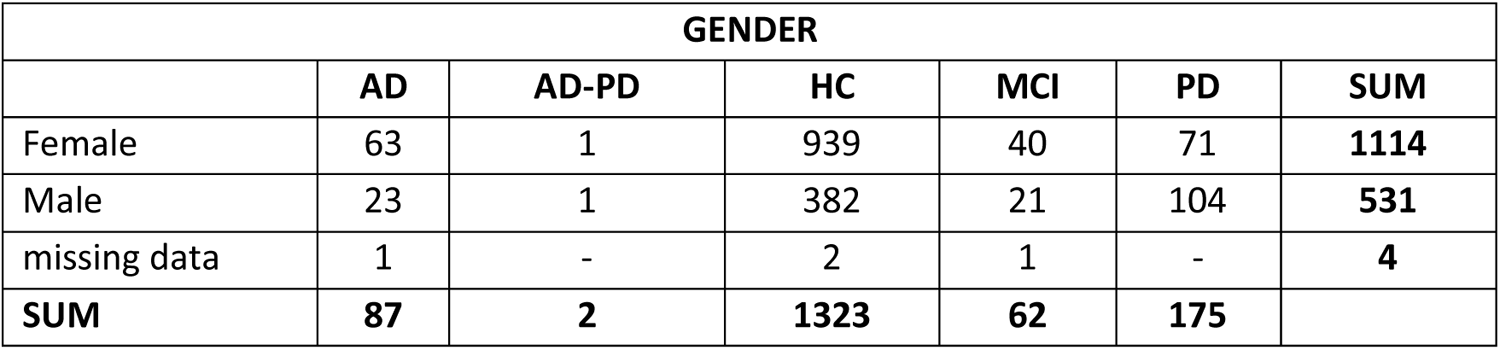
Distribution of EWA-DB according to gender.

When recruiting participants in clinical samples, or in the sample of healthy controls, the predefined inclusion criteria (see Inclusion and exclusion criteria) were followed. In several cases, neurologists or psychiatrists gave the diagnosis of AD, PD or MCI (or the diagnosis was refuted, and the participants were marked as healthy), but within the inclusion criteria the participants did not meet some conditions (e.g., MoCA scores). In such cases, the participants were not excluded and are still part of the database. If we were to consider precisely defined inclusion and exclusion criteria, the size of the database would be 1122 speakers. Within the database, for each speaker, there is information about meeting / not meeting the inclusion criteria. The distribution of the database in terms of age, education and gender while meeting the inclusion criteria is shown in Tables 4, 5 and 6.

**Table 4.** Distribution of EWA-DB according to age when meeting the inclusion criteria.

| **AGE** |  |  |  |  |  |  |
| --- | --- | --- | --- | --- | --- | --- |
|  | **AD** | **AD-PD** | **HC** | **MCI** | **PD** | **SUM** |
| <50 | 0 | 0 | 61 | 0 | 13 | **74** |
| 50-59 | 0 | 0 | 286 | 0 | 22 | **308** |
| 60-69 | 3 | 0 | 338 | 5 | 61 | **407** |
| 70-79 | 10 | 0 | 176 | 13 | 53 | **252** |
| >=80 | 14 | 0 | 34 | 12 | 20 | **80** |
| missing data | 0 | 0 | 1 | 0 | 0 | **1** |
| **SUM** | **27** | **0** | **896** | **30** | **169** |  |

**Table 5.**
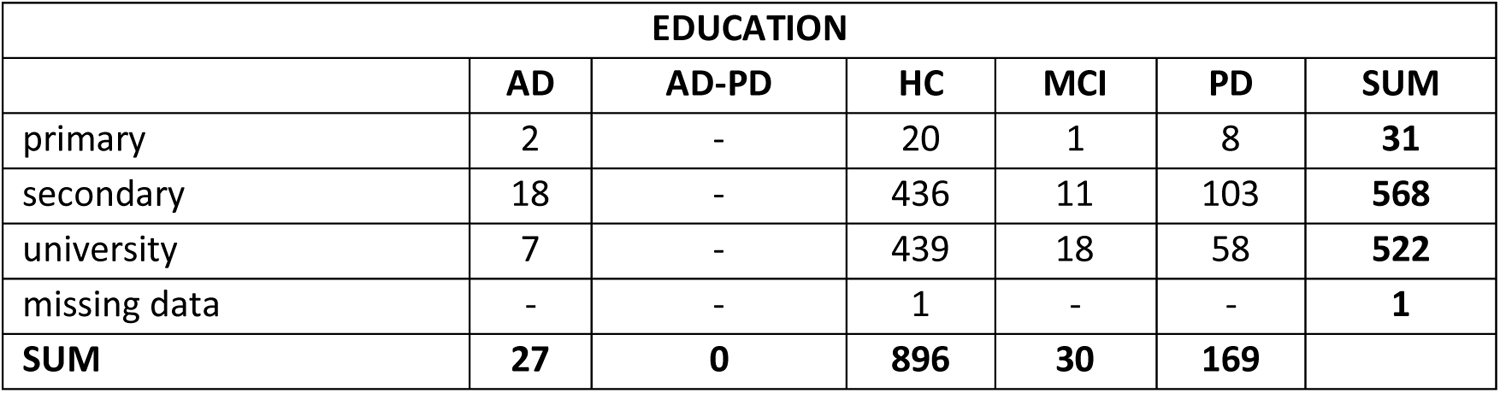
Distribution of EWA-DB according to education when meeting the inclusion criteria.

| **EDUCATION** |  |  |  |  |  |  |
| --- | --- | --- | --- | --- | --- | --- |
|  | **AD** | **AD-PD** | **HC** | **MCI** | **PD** | **SUM** |
| primary | 2 | - | 20 | 1 | 8 | **31** |
| secondary | 18 | - | 436 | 11 | 103 | **568** |
| university | 7 | - | 439 | 18 | 58 | **522** |
| missing data | - | - | 1 | - | - | **1** |
| **SUM** | **27** | **0** | **896** | **30** | **169** |  |

**Table 6.** Distribution of EWA-DB according to gender when meeting the inclusion criteria.

| **GENDER** |  |  |  |  |  |  |
| --- | --- | --- | --- | --- | --- | --- |
|  | **AD** | **AD-PD** | **HC** | **MCI** | **PD** | **SUM** |
| Female | 20 | - | 647 | 19 | 66 | **752** |
| Male | 7 | - | 248 | 11 | 103 | **369** |
| missing data | - | - | 1 | - | - | **1** |
| **SUM** | **27** | **0** | **896** | **30** | **169** |  |

Since not all participants agreed to the publication of their audio recordings, out of the total number of 1649 speakers, the database contains the recordings of 1003 speakers, i.e., those from whom we have written consent.

However, all 1649 speakers gave written consent to the processing of their recordings. The recordings of all participants were transcribed using automated speech recognition. Subsequently, the transcriptions were corrected and annotated by trained annotators (see Annotation). ASR transcription is available for every speaker (N = 1649), even for those speakers who did not give written consent to the publication of the audio recording. Manually annotated transcripts are available for 1502 speakers – for every speaker from the clinical sample and for the majority of speakers from the control sample.

### Structure of EWA-DB

All files of the database are contained in a main folder named EWA-DB. The structure of this folder is depicted in Fig. 2.

**Fig. 2.**
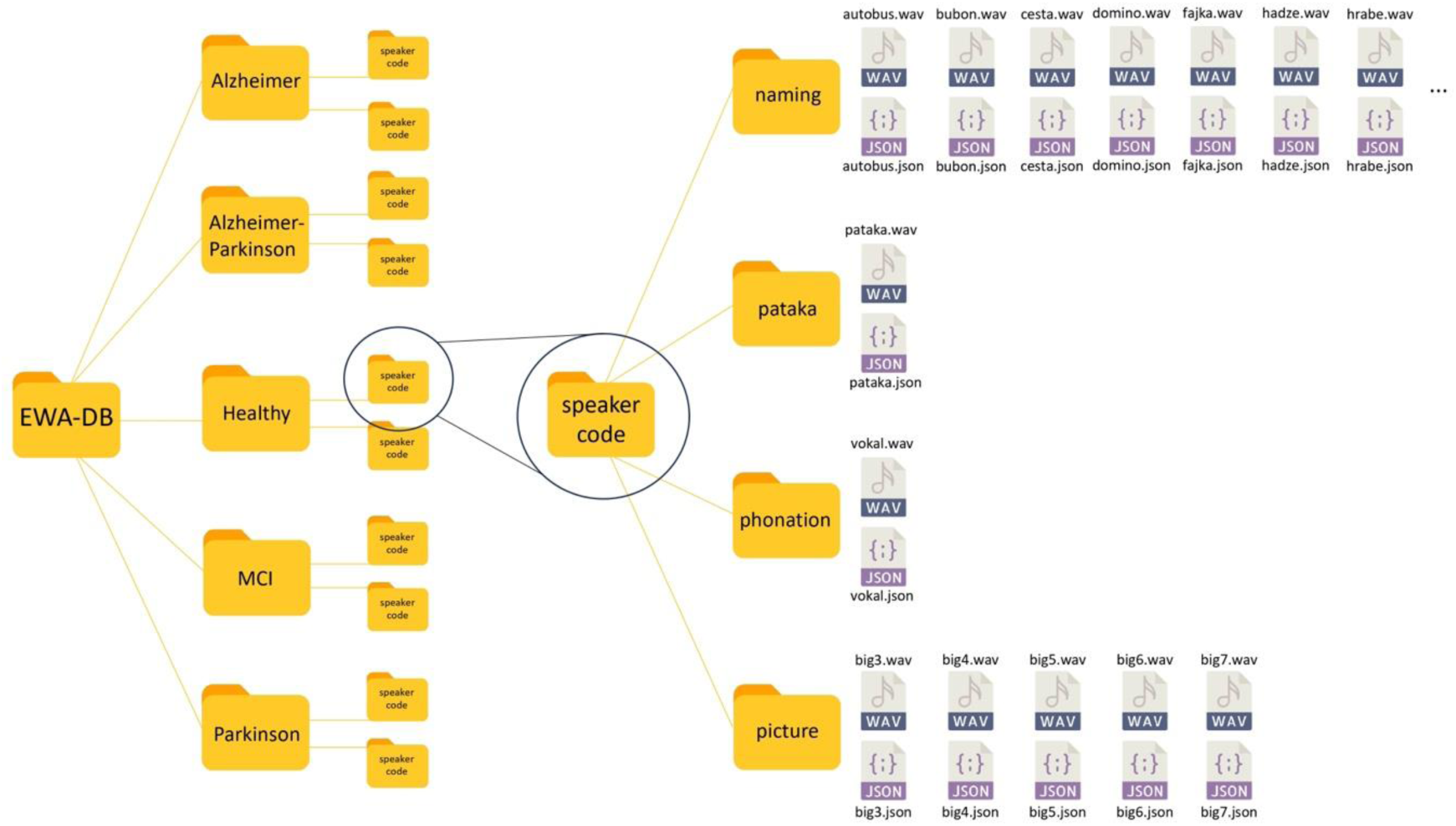
Structure of EWA-DB.

After opening EWA-DB, additional folders will be displayed - one for each clinical group and one for the group of healthy controls. In each group of participants, there are subsequent folders for individual speakers, which are marked with randomly generated codes. After opening the folder of one speaker, folders for individual language tasks will be displayed - that is, separate folders for phonation, diadochokinesis (pataka), naming and picture description.

If the speaker gave consent the folders contain recordings as WAV files and additional JSON files (UTF-8 character encoding) containing ASR transcription, annotation, speaker information, Trill Embedding, acoustic features measured via Neurospeech and OpenSmile (Gemaps set). If the consent to publish recordings was not given, only a JSON file is provided.

The folder *phonation* contains the task of sustained vowel phonation.

The folder *pataka* contains the task for diadochokinesis.

The folder *naming* contains several files – one WAV file and one JSON file for every picture used in the object and action naming task. These files are marked with the correct naming response in the Slovak language. The name of each file is in Slovak. In table 7 we present the target items of object and action naming in English. The database includes a file (EWA- DB_paper_preprint_1.pdf) listing Slovak target items with corresponding English translation.

**Table 7.**
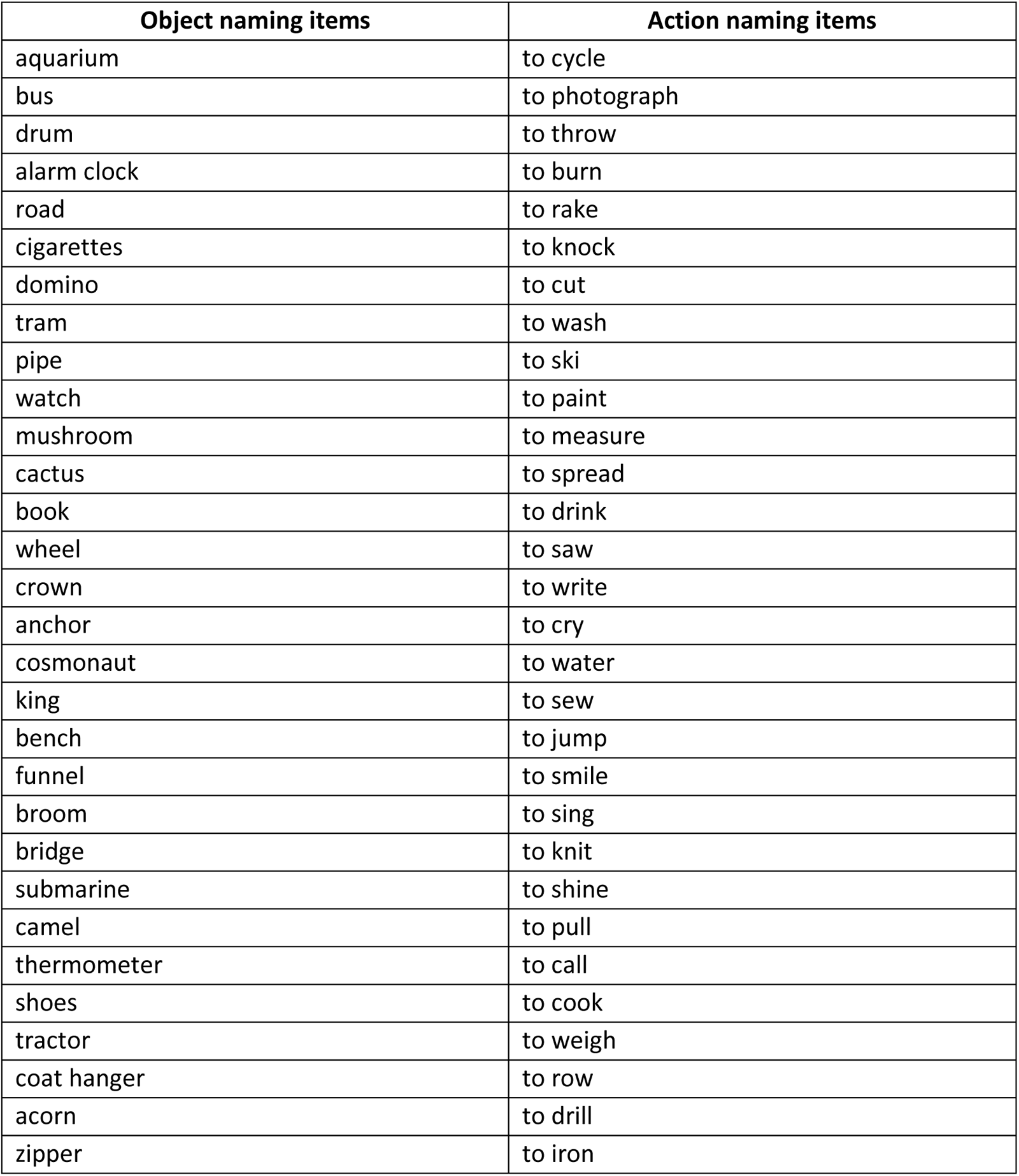
Corresponding English terms for the Slovak naming items.

The folder *picture description* contains the descriptions of three complex pictures named *big3*, *big 4* and *big 5* and two simple pictures named *big 6* and *big 7*.

## Technical Validation

### Characteristics of EWA-DB in terms of age, gender, education and MoCA score

The database contains a complete sample of 1122 participants meeting the inclusion criteria. In Table 8, for each group of patients with AD, MCI, PD and the group of healthy controls, the summary numbers of participants, the mean age and the mean MoCA score by education and gender are presented.

**Table 8.** Samples composition of the EWA database.

|  | AD |  | MCI |  | PD |  | HC |  |
| --- | --- | --- | --- | --- | --- | --- | --- | --- |
| Participants | Male | Female | Male | Female | Male | Female | Male | Female |
| Primary | 1 | 1 | - | 1 | 2 | 6 | 6 | 14 |
| Secondary | 4 | 14 | 3 | 8 | 59 | 44 | 115 | 322 |
| University | 2 | 5 | 8 | 10 | 42 | 16 | 127 | 312 |
| **Total participants** | **7** | **20** | **11** | **19** | **103** | **66** | **248** | **648** |
| AGE average | Male | Female | Male | Female | Male | Female | Male | Female |
| Primary | 71.0 | 83.0 | - | 80.0 | 57.0 | 72.5 | 68.0 | 69.8 |
| Secondary | 73.8 | 81.1 | 72.7 | 77.1 | 64.7 | 70.3 | 61.6 | 69.6 |
| University | 78.5 | 81.4 | 75.6 | 76.3 | 68.7 | 63.4 | 62.8 | 60.8 |
| **Age total average** | **74.7** | **81.3** | **74.8** | **76.8** | **66.2** | **68.8** | **62.4** | **65.4** |
| MOCA average | Male | Female | Male | Female | Male | Female | Male | Female |
| Primary | 19.0 | 20.0 | - | 25.0 | 28.0 | 26.3 | 28.0 | 27.1 |
| Secondary | 21.0 | 19.8 | 23.7 | 23.8 | 25.6 | 25.6 | 27.5 | 27.8 |
| University | 19.5 | 20.8 | 23.9 | 23.7 | 25.8 | 26.4 | 27.9 | 28.1 |
| **MOCA average** | **20.3** | **20.1** | **23.8** | **23.8** | **25.7** | **25.9** | **27.8** | **27.9** |

As shown in Table 8, the smallest number of participants is in the group of patients with AD (N=27). The total number of MCI patients is (N=30). There are fewer males than females in both the AD and MCI patient groups. The total number of patients with PD is (N=169) and unlike the other groups, the number of men (N=103) is higher than the number of women (N=66). In our sample, the average age of patients with AD and MCI increases with higher education in men. On the contrary, in women, the average age decreases with education. In the case of primary and secondary education, the average age is higher for women than for men in all groups. There are no notable differences in mean MoCA scores within genders according to education.

**Fig. 3.**
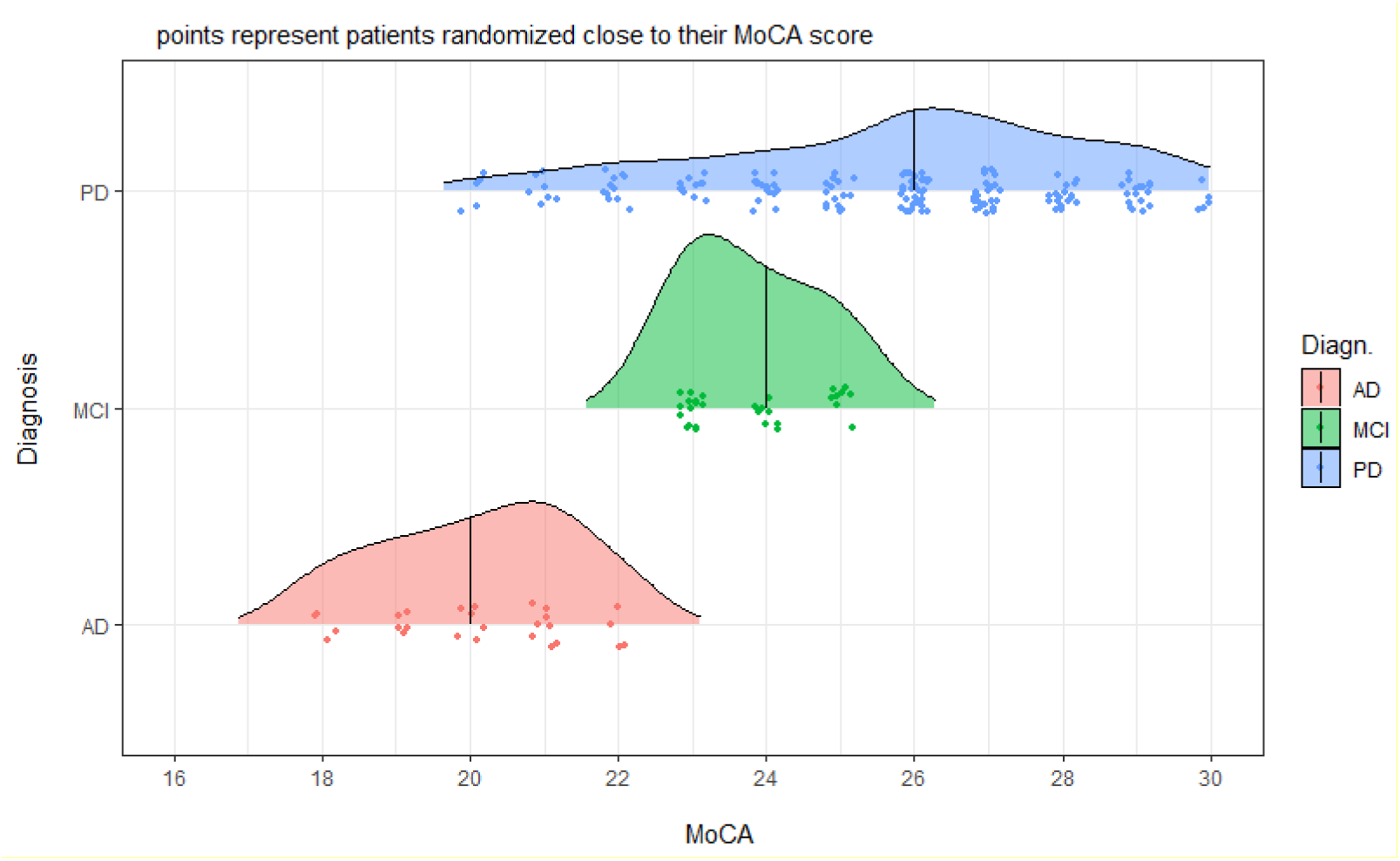
Density distribution plots for clinical samples AD, MCI, and PD according to their MoCA score.

The patient density graph for AD patients by MoCA score in Fig. 3 shows that the median of MoCA score is 20 and the largest number of AD patients have a MoCA score of 21 above the median. In MCI patients, on the contrary, the MoCA median is 24, but the largest number of patients have a MoCA score below 23. This suggests that there is a natural transition of MCI patients to AD according to MoCA scores. The density distribution plot of PD patients in Fig. 5 shows that the median of MoCA score is 26 and the largest number of patients have a MoCA score of 26. The density distribution of PD patients is more elongated towards lower MoCA score values of 20.

**Fig. 4.**
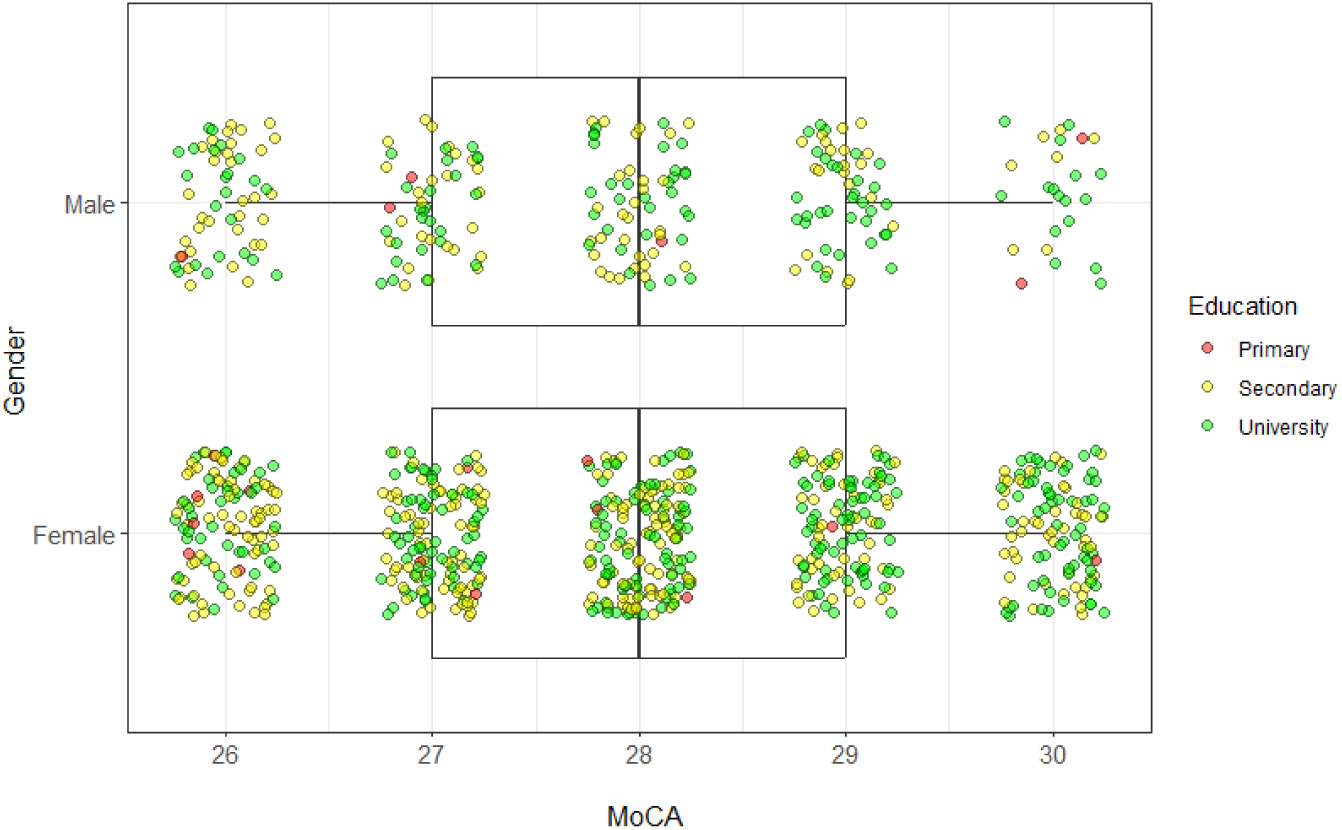
Density graph of healthy control distribution according to MoCA, gender and education.

Fig. shows the distribution of the healthy control group according to MoCA scores, gender, and education by visual evaluation using the boxplot function of the statistical software R. The coloured points represent healthy participants according to education (primary, secondary, and university), who are randomly distributed near their MoCA score in both axes. Separately, the distribution by gender for men and women is shown. From the given distribution, it follows that women are evenly distributed for MoCA scores of 26-30 with a median of 28. For the control group of healthy men, the distribution with fewer participants is similar to the distribution in women, with a median MoCA of the same value (28) and the same interquartile range.

**Fig. 5.**
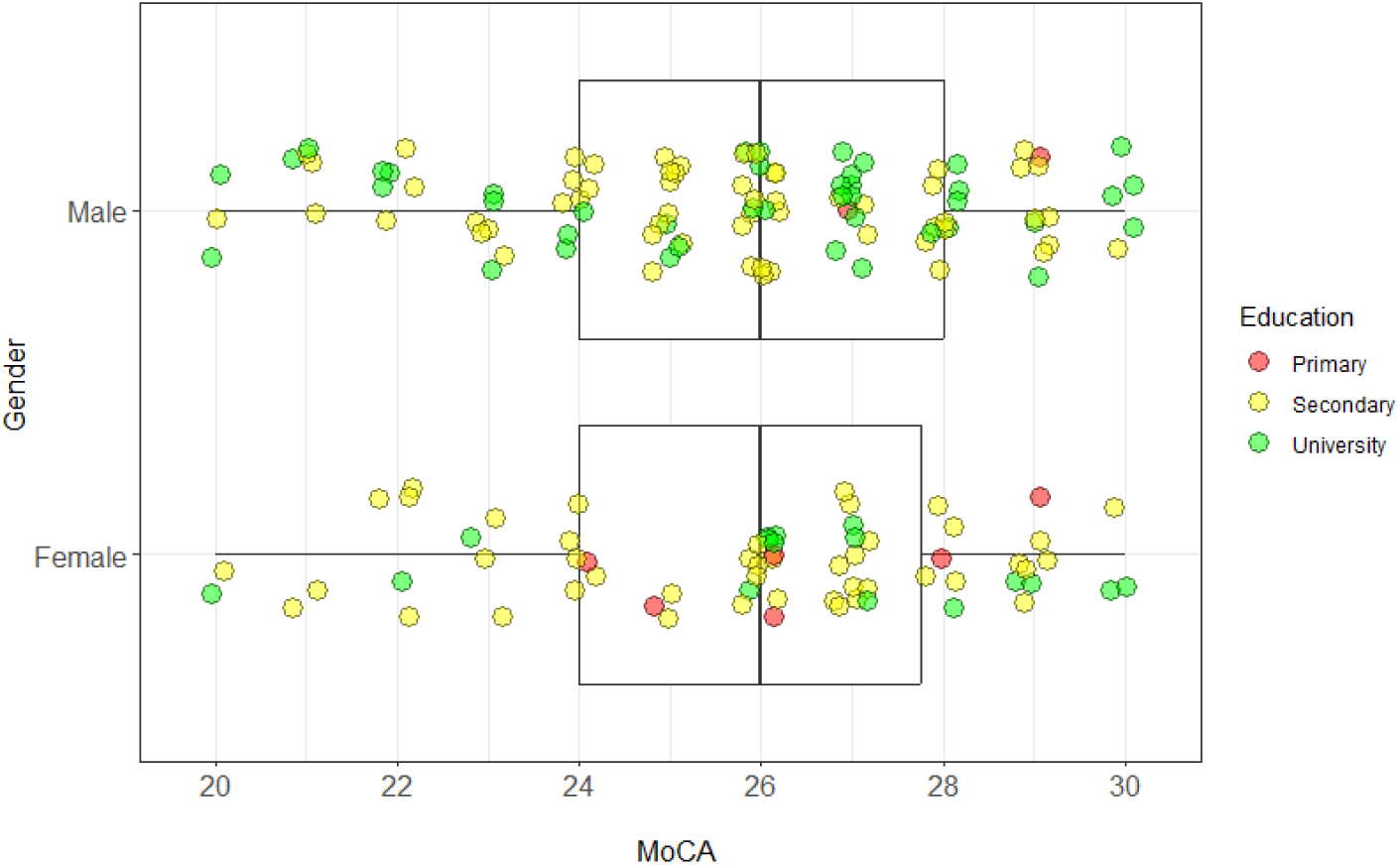
Density graph of PD patient distribution according to MoCA, gender and education.

Fig. 5 shows a visual representation of the distribution of Parkinson’s disease patients according to MoCA scores, gender, and education. Coloured points represent PD patients according to education (primary, secondary, and university), who are distributed near their MoCA scores. This distribution is presented separately for male and female PD patients. From the given distribution the MoCA median is the same for men and women (26) and the distribution of the quartiles is the same, with the lower quartile being longer with a MoCA score range of 24-26. Among women with PD, secondary education is noticeably dominant.

**Fig. 6.**
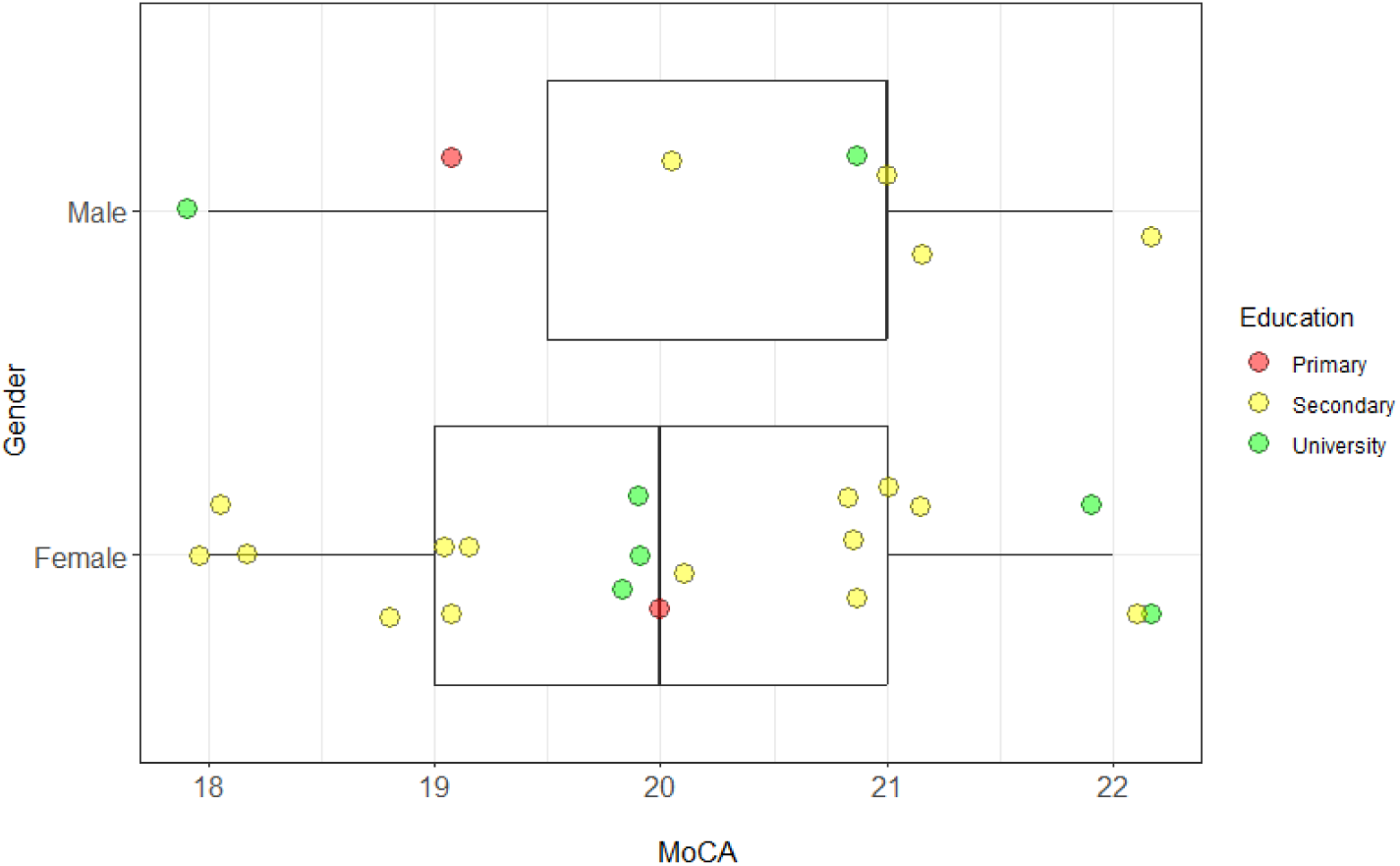
Density graph of AD patient distribution according to MoCA, gender and education.

**Fig. 7.**
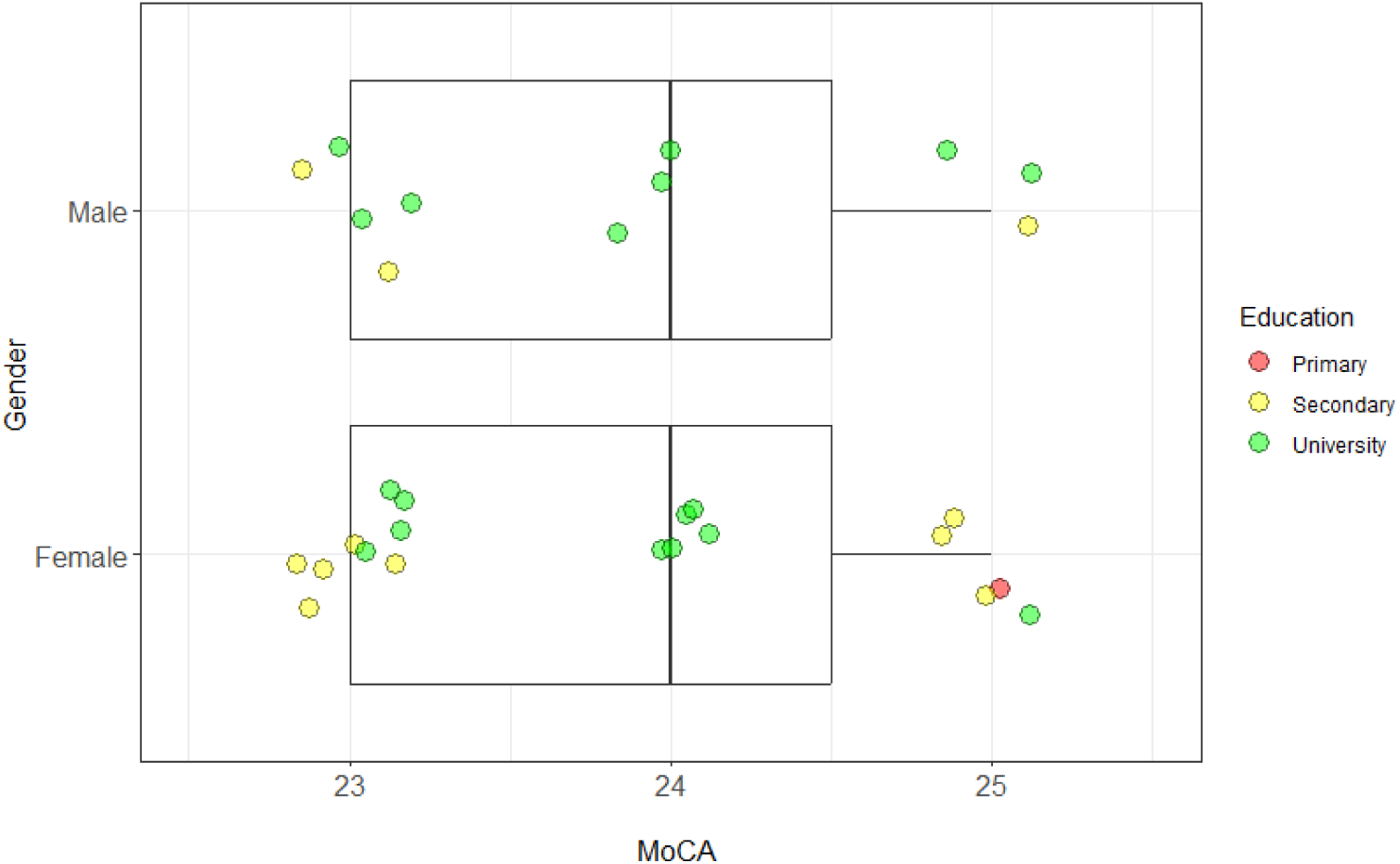
Density graph of MCI patient distribution according to MoCA, gender and education.

The number of patients with Alzheimer’s disease (N=26) and MCI (N=30) is small, so the representation of patients can be directly observed in Fig. 6 and Fig. 7. There is a low representation of men with AD, with the MoCA median (21) being higher than the MoCA median in women (20). Most of AD patients have secondary education. For MCI patients, the MoCA median (24) is the same for men and women, with the highest number of MCI patients with a university degree.

### Quality control of recordings and transcripts

During the EWA project, recordings were made at various clinical sites. Especially at the beginning of the project, it occurred that during the recording there were disturbing noises and sounds in the background, such as the siren of an ambulance, noise from the corridor, interruption by medical personnel, knocking on the door, or the administrator taking notes near the microphone during the testing procedure. Disturbing moments like these can have an impact on sound parameters and affect the process and results of machine learning. For this reason, a subjective evaluation of the acoustic quality of the recordings was performed. Every recording that was considered acoustically unsatisfactory was marked as low quality. Information about the quality of each recording is also provided in the JSON file.

The next phase of quality control took place in the transcription process. Automatic speech recognition was used to create transcriptions from speech recordings. These transcripts were consequently checked and corrected by trained annotators, most of whom were speech and language pathology students who already gained basic experience with creating transcriptions during their studies. Each annotator took part in a detailed training at the Institute of Informatics of Slovak Academy of Sciences. As part of this training, the annotators learned how to properly check, correct, and annotate transcripts and how to work with the Transcriber 1.5.1 program^88^. At first, the work of the annotators was checked by their supervisor, a speech and language pathologist, until the conclusion was made, that the annotators had adopted the established rules for annotation and could work independently. Annotators received transcripts as TRS files and corresponding recordings as WAV files. In addition to entering tags (see Annotation), the task of each annotator was to check the transcription and, in case of inconsistencies, to correct them. When annotating the picture description tasks, annotators divided the transcription into sentences, following intonation, semantics, and syntax. Out of the total number of 1649 transcripts, 1502 is manually annotated. If the annotation is available, it is included in the JSON file.

## Data Availability

All data presented in the present work are publicly available at ELRA/ELDA under the name EWA-DB Early Warning of Alzheimer speech database. (2023)

https://catalog.elra.info/en-us/repository/browse/ELRA-S0489/

## Acknowledgements

This study was created in relation to the project EWA - Early Warning of Alzheimer (ITMS2014+ : 313022V631), which was funded by the European Regional Development Fund and the project ALOIS - Diagnosis of Alzheimer’s disease from speech using artificial intelligence and social robotics (APVV-21-0373), which was funded by the Slovak Research and Development Agency.

## Author contribution

Milan Rusko, Alfréd Zimmermann, and Eugen Ružický are the leaders of the research teams of the members of the consortium, they participated in the organization of the creation of the database and the collection of recordings.

Matej Škorvánek and Petra Brandoburová were the scientific and clinical guarantors of the project, the main authors of the methodology, as well as the organizers and supervisors of the recording process.

Richard Malaschitz and Marián Trnka provided the necessary technical solutions, designed the data collection and storage system, as well as data processing into the definitive form of the database.

Róbert Sabo and Viktória Kevická were responsible for manual annotation of transcripts, namely recruiting, training, and supervising the work of annotators.

## Competing interests

All authors declare that they have no conflicts of interest.

## Code Availability

No custom code has been used.

## Notes

### Competing Interest Statement

The authors have declared no competing interest.

### Author Declarations

Ethics committee of Bratislava self-governing region (Bratislavský samosprávny kraj) gave ethical approval for this work.

